# Clinically actionable pharmacogenomic landscape of antidepressants and antipsychotics in Qatar: A population-based cohort study

**DOI:** 10.1101/2023.09.27.23296201

**Authors:** Dinesh Velayutham, Kholoud Bastaki, the Qatar Genome Program Research Consortium, Suhaila Ghuloum, Muhammad Waqar Azeem, Munir Pirmohamed, Puthen Veettil Jithesh

**Affiliations:** College of Health & Life Sciences, Hamad Bin Khalifa University, Doha, Qatar; Qatar University, Doha, Qatar; Department of Psychiatry, Hamad Medical Corporation, Doha, Qatar; Department of Psychiatry, Sidra Medicine, Doha, Qatar; Institute of Systems, Molecular and Integrative Biology, University of Liverpool, United Kingdom

## Abstract

Antidepressants and antipsychotics, commonly prescribed worldwide, exhibit significant variability in patient responses. Genetic variations contribute substantially to this variability, offering the potential for predicting individual responses through pharmacogenetic testing. Clinical guidelines provided by organizations like the Clinical Pharmacogenetic Implementation Consortium (CPIC) and the Dutch Pharmacogenetic Working Group (DPWG) facilitate the application of pharmacogenomics in clinical settings. Understanding the prevalence of actionable genetic variants and their associated response phenotypes is vital for effective pharmacogenomic integration, especially in under-served populations.

In this study, we analyzed the frequency distribution of actionable genetic variants in 6045 adult Qataris recruited by the Qatar Biobank, whose genomes were sequenced as part of the Qatar Genome Program, focusing on genes (CYP2C19, CYP2D6, CYP2B6, and CYP3A4) influencing psychotropic medication responses. Haplotypes and diplotypes were generated from 490 alleles, and metabolizer phenotypes were predicted based on established guidelines.

Our findings revealed that over 52% of Qatari individuals may possess actionable metabolizer phenotypes associated with CYP2C19, CYP2B6, and CYP2D6, impacting their response to tricyclic antidepressants. The frequency of actionable genetic variants for serotonin reuptake inhibitors ranged from approximately 2% to 51%, and for antipsychotics, it ranged from 0.1% to 32%, based on genetic variations in CYP3A4 and CYP2D6.

This study underscores the importance of assessing these genetic variants, particularly concerning commonly prescribed antidepressants like escitalopram and amitriptyline in Qatar. Implementing pharmacogenetic testing based on these findings can potentially enhance patient outcomes in psychiatric medication management.

## Introduction

Antidepressants and antipsychotics are widely prescribed in Qatar^1,2^ and worldwide, however, response is highly variable. Genetic variants contribute to inter-individual variability, and pharmacogenetic testing provides a means to predict potential non-response. Guidelines for the clinical implementation of pharmacogenetic tests for several antidepressants and antipsychotics have been provided by the Clinical Pharmacogenetic Implementation Consortium (CPIC) and the Dutch Pharmacogenetic Working Group (DPWG). Understanding the distribution of clinically actionable genotypes and their predicted response phenotypes is essential in facilitating clinical implementation of pharmacogenomics in different settings including under-served populations. Therefore, we have examined the frequency distribution of actionable genetic variants in the Qatari population from the Middle East.

## Methods

Following approval from the QBB Institutional Review Board, we have studied an observational cohort of 6218 adult Qataris recruited to the Qatar Biobank (QBB) between 11 December 2012 and 9 June 2016. Their genomes were sequenced as part of the pilot phase of the Qatar Genome Program (QGP) and 6045 were taken forward following quality analysis of the whole genome sequencing data. Further details of the cohort and genome data processing have been presented previously^3^.

We analyzed genes coding for metabolizing enzymes (*CYP2C19, CYP2D6, CYP2B6* and *CYP3A4)* that significantly affect response to psychotropics and have guidelines for clinical interpretation^4-6^. We extracted 490 alleles to generate haplotypes and diplotypes and predicted the associated metabolizer phenotypes based on PharmVar (https://www.pharmvar.org/) and CPIC translation tables (https://cpicpgx.org/genes-drugs/). Clinically actionable pharmacogenomic variance was defined based on clinical guidelines from CPIC and DPWG suggesting a course of action, such as change in dosage or prescription of an alternate drug. Custom Python scripts and Cyrius (https://github.com/Illumina/Cyrius) were used for the analyses. For statistical presentation of results, we used absolute numbers and percentages.

## Results

Genome sequencing data from 6045 Qataris showed that more than 52% of the individuals may have actionable metabolizer phenotype associated diplotypes in *CYP2C19, CYP2B6* and *CYP2D6* affecting response to tricyclic antidepressants. For the serotonin reuptake inhibitors, the frequency varied from ∼2-51%, while for antipsychotics, the range was 0.1-32% based on genetic variation in *CYP3A4* and *CYP2D6* (**Table)**.

**Table:** The distribution of actionable phenotypes predicted from diplotypes affecting response to psychotropics and requiring alteration of dosage or alternate prescription in the Qatari population from 6045 whole genomes.

| Class of drug | Drugs (Pharmacogenes) | Metabolizer Phenotype: # (%) | Major Clinical Action (For detailed recommendation, please see the references) | Total Actionable # (%) |
| --- | --- | --- | --- | --- |
| Antidepressants-selective serotonin reuptake inhibitors (SSRIs)<sup>#</sup> | Citalopram<br>Escitalopram (CYP2C19) | UM: 394 (6.5%)<br>PM: 111 (1.8%) | Use an alternate antidepressant not mainly metabolized by CYP2C19. If these are the best options, titrate to higher maintenance dose, if required. | 3075 (50.9%) |
|  |  | RM: 1520 (25.1%) | Titrate to higher maintenance dose or use alternate antidepressant not mainly metabolized by CYP2C19, if required. |  |
|  |  | IM: 1050 (17.4%) | Use a slower titration schedule and lower maintenance dose than normal metabolizers. |  |
|  | Sertraline (CYP2C19 + CYP2B6) | C19 UM/RM + B6 UM/RM: 77 (1.3%) | If not responding with standard dose, titrate to a higher dose or switch to another antidepressant not mainly metabolized by CYP2C19/CYP2B6 | 2566 (42.4%) |
|  |  | C19 IM + B6 NM: 434 (7.2 %)<br>C19 NM + B6 IM: 1127 (18.6%)<br>C19 IND + B6 IM: 3 (0.05%)<br>C19 IM + B6 IND: 58 (0.96%)<br>C19 IM + B6 IM: 420 (6.9%) | Use slower titration schedule and lower maintenance dose |  |
|  |  | C19 PM + B6 NM:42 (0.69%)<br>C19 PM + B6 IM: 47 (0.78%)<br>C19 PM + B6 IND: 8 (0.13%)<br>C19 PM + B6 UM/RM: 3 (0.05%) | Use lower starting dose, slower titration schedule and 50% reduction of usual maintenance dose as compared to C19 NM or select another antidepressant not metabolized by C19 |  |
|  |  | C19 NM + B6 PM:252 (4.2%)<br>C19 IND + B6 PM 2 (0.03%) | Use lower starting dose, slower titration schedule and 25% reduction of standard maintenance dose as compared to B6 NM or select another antidepressant not metabolized by B6 |  |
|  |  | C19 IM + B6 PM: 84 (1.4%) | Use lower starting dose, slower titration schedule and 50% reduction of standard maintenance dose as compared to B6 NM |  |
|  |  | C19 PM + B6 PM: 9 (0.15%) | Select alternative antidepressant not metabolized by C19/B6. |  |
|  | Fluvoxamine (CYP2D6) | PM: 102 (1.7%) | Use 25-50% lower starting dose and slower titration compared to CYP2D6 NM or consider alternative antidepressant not mainly metabolized by CYP2D6 | 102 (1.7%) |
|  | Paroxetine (CYP2D6) | UM: 523 (8.6%) | Use alternate drug not metabolized by CYP2D6 | 1931 (31.9%) |
|  |  | PM: 102 (1.7%) | Use 50% reduction in recommended starting dose, slower titration, and 50% lower maintenance dose compared to CYP2D6 NM |  |
|  |  | IM: 1306 (21.6%) | Use lower starting dose and slower titration compared to CYP2D6 NM |  |
| Antidepressants-serotonin & norepinephrine reuptake inhibitors SNRIs<sup>#</sup> | Venlafaxine (CYP2D6) | PM: 102 (1.7%) | Use an alternate antidepressant not mainly metabolized by CYP2D6 | 102 (1.7%) |
| Antidepressants-SSRI-like serotonin modulators<sup>#</sup> | Vortioxetine (CYP2D6) | UM: 523 (8.6%) | Use an alternate antidepressant not mainly metabolized by CYP2D6. If required, increase maintenance dose by 50% | 625 (10.3%) |
|  |  | PM: 102 (1.7%) | Initiate 50% of starting dose and titrate to maximum recommended dose or use an alternate antidepressant not mainly metabolized by CYP2D6 |  |
| Antidepressants-TCAs<sup>▽</sup> | Amitriptyline<br>clomipramine<br>doxepin<br>imipramine<br>trimipramine<br>(CYP2C19 + CYP2D6) | C19 UM/RM + D6 UM/RM: 186 (3.1%)<br>C19 UM/RM + D6 PM: 33 (0.5%)<br>C19 IM + D6 UM/RM: 64 (1.1%)<br>C19 PM + D6 UM/RM: 5 (0.1%)<br>C19 PM + D6 IM: 30 (0.5%)<br>C19 PM + D6 PM 1 (0.01%)<br>C19 NM + D6 UM/RM: 268 (4.4%) | Avoid TCA use | 3172 (52.5%) |
|  |  | C19 UM/RM + D6 NM: 1181 (19.5%)<br>C19 UM/RM + D6 IM: 409 (6.8%) | Consider alternative drug not metabolized by C19 |  |
|  |  | C19 PM + D6 NM: 60 (1%)<br>C19 NM + D6 PM: 55 (0.9%)<br>C19 IM + D6 PM: 13 (0.2%) | Avoid amitriptyline, but if warranted, use 50% of the starting dose |  |
|  |  | C19 NM + D6 IM: 603 (10%)<br>C19 IM + D6 IM: 264 (4.4%) | Consider 25% reduction in starting dose |  |
| Antipsychotics<sup>∞</sup> | Quetiapine (CYP3A4) | PM: 6 (0.1%) | For depression, use alternative.<br>For other indications, use 30% of the normal dose. | 6 (0.1%) |
|  | Aripiprazole (CYP2D6) | PM: 102 (1.7%) | Use 68-75% of the normal maximum dose of aripiprazole. | 102 (1.7%) |
|  | Brexipiprazole (CYP2D6) | PM: 102 (1.7%) | Use half the normal dose | 102 (1.7%) |
|  | Risperidone (CYP2D6) | UM: 523 (8.6%) | Use an alternative or titrate the dose according to the maximum dose for the active metabolite | 625 (10.3%) |
|  |  | PM: 102 (1.7%) | Use 67% of the normal dose. Reduce the dose further to 50% of the normal dose, if adverse reactions occur |  |
|  | Haloperidol Risperidone (CYP2D6) | UM: 523 (8.6%) | Use 1.5 times the normal dose or an alternative drug | 625 (10.3%) |
|  |  | PM: 102 (1.7%) | Use 60% of the normal dose. |  |
|  | Pimozide (CYP2D6) | PM: 102 (1.7%) | Use no more than 50% of the normal maximum dose | 1408 (23.3%) |
|  |  | IM: 1306 (21.6%) | Use no more than 80% of the normal maximum dose |  |
|  | Zuclopenthixol (CYP2D6) | PM: 102 (1.7%) | Use 50% of the normal dose | 1931 (31.9%) |
|  |  | IM: 1306 (21.6%) | Use 75% of the normal dose |  |
|  |  | UM: 523 (8.6%) | If not effective, increase dose upto 1.5x normal dose |  |

Metabolizer status - UM: ultrarapid; RM: rapid; PM: poor; IM: intermediate; NM: normal; IND: indeterminate

## Discussion

We predicted the distribution of CYP2B6, CYP2C19, CYP2D6 and CYP3A4 metabolizer phenotypes in 6045 Qatari individuals from multiple variants in each of these genes, including copy number changes and based on the latest guidelines for modifying the prescriptions of antidepressants and antipsychotics, identified the distribution of actionable frequencies in the Qatari population. For example, just over 50% of the population may need alteration in prescription of escitalopram and amitriptyline. Since both these drugs are among the most commonly prescribed antidepressants in Qatari patients, our results highlight the importance of assessing the role of these variants, and their implementation into clinical practice to improve patient outcomes.

A limitation of this study is the prediction of phenotypes based on existing literature from other populations. Although it can be safely assumed that the effects of the diplotypes should be similar in different populations, population-specific rare variants were not included in this study.

## Data Availability

The informed consent given by the study participants does not cover posting of participant level phenotype and genotype data of Qatar Biobank/Qatar Genome Project in public databases. However, access to QBB/QGP data can be obtained through an established ISO-certified process by submitting a project request at
https://www.qatarbiobank.org.qa/research/how-apply which is subject to approval by the QBB IRB committee.

## Acknowledgements

We thank the QBB and QGP for providing in-kind funding through the access to the whole genome sequencing data for this study as part of the QGP Research Consortium. The funders had no role in the interpretation of data. We thank Mohammed Abuhaliqa, Ikhlak Ahmed, Mohammed ElAnbari and Najeeb Syed, Anjanarani N and Mashael Alshafai for their involvement in the preliminary analysis or management.

MP receives research funding from various organisations including the UK MRC and NIHR. He has also received partnership funding for the following: MRC Clinical Pharmacology Training Scheme (co-funded by MRC and Roche, UCB, Eli Lilly and Novartis); a PhD studentship jointly funded by EPSRC and Astra Zeneca; and grant funding from Vistagen Therapeutics. He has also unrestricted educational grant support for the UK Pharmacogenetics and Stratified Medicine Network from Bristol-Myers Squibb and UCB. He has developed an HLA genotyping panel with MC Diagnostics, but does not benefit financially from this. He is part of the IMI Consortium ARDAT (www.ardat.org). MP is also Vice Chair of the Qatar Precision Medicine Initiative International Scientific Advisory Committee.

## Supplementary Information

**The Qatar Genome Program Research Consortium**

(names in alphabetical order)

Rania Abdel-latif ^4^, Tariq Abu Saqri ^2^, Tariq Abu Zaid ^2^, Nahla Afifi ^8^, Rashid Al-Ali ^2^, Souhaila Al-Khodor ^2^, Wadha Al-Muftah ^4^, Yasser Al-Sarraj ^4^, Omar Albagha ^1,9^, Eiman Alkhayat ^8^, Fatima Alkuwari ^4^, Hakeem Almabrazi ^2^, Mashael Alshafai ^5^, Asmaa Althani ^4,8^, Muhammad Alvi ^4^, Ramin Badii ^3^, Radja Badji ^4^, Lotfi Chouchane ^10^, Dima Darwish ^4^, Ahmed El Khouly ^2^, Maryem Ennaifar ^4^, Xavier Estivill ^6^, Tasnim Fadl ^4^, Khalid Fakhro ^2,10^, Eleni Fethnou ^8^, Mehshad Hamza ^2^, Said I. Ismail ^4^, Puthen V. Jithesh ^1^, Mohammedhusen Khatib ^2^, Wei Liu ^2^, Stephan Lorenz ^2^, Hamdi Mbarek ^4^, Younes Mokrab ^2^, Tushar Pathare ^2^, Shafeeq Poolat ^2^, Fatima Qafoud ^8^, Fazulur Rehaman Vempalli ^2^, Chadi Saad ^4^, Karsten Suhre ^10,11^, Najeeb Syed ^2^, Zohreh Tatari ^2^, Ramzi Temanni ^2^, Sara Tomei ^2^, Heba Yasin ^4^

^1^ College of Health & Life Sciences, Hamad Bin Khalifa University, Doha, Qatar

^2^ Research Branch, Sidra Medicine, Doha, Qatar

^3^ Hamad Medical Corporation, Doha, Qatar

^4^ Qatar Genome Program, Qatar Foundation Research Development and Innovation, Doha, Qatar

^5^ Department of Biomedical Sciences, College of Health Sciences, Qatar University, Doha, Qatar

^6^ Quantitative Genomics Laboratories, Barcelona, Catalonia, Spain

^8^ Qatar Biobank for Medical Research, Qatar Foundation, Doha, Qatar

^9^ Institute of Genetics and Molecular Medicine, University of Edinburgh, Edinburgh, UK

^10^ Weill Cornell Medicine-Qatar, Doha, Qatar

^11^ Weill Cornell Medicine, New York, NY, USA

## Notes

### Competing Interest Statement

Munir Pirmohamed (MP) receives research funding from various organisations including the UK MRC and NIHR. He has also received partnership funding for the following: MRC Clinical Pharmacology Training Scheme (co-funded by MRC and Roche, UCB, Eli Lilly and Novartis); a PhD studentship jointly funded by EPSRC and Astra Zeneca; and grant funding from Vistagen Therapeutics. He has also unrestricted educational grant support for the UK Pharmacogenetics and Stratified Medicine Network from Bristol-Myers Squibb and UCB. He has developed an HLA genotyping panel with MC Diagnostics, but does not benefit financially from this. He is part of the IMI Consortium ARDAT (www.ardat.org). MP is also Vice Chair of the Qatar Precision Medicine Initiative International Scientific Advisory Committee.

### Funding Statement

PVJ is supported by faculty funding from the College of Health & Life Sciences,
HBKU and the in-kind funding for access to data from Qatar Biobank and Qatar Genome Program, Qatar Foundation for Education, Science and Community Development. Funders had no role in the design of the study and collection, analysis,
and interpretation of data and in writing the manuscript.

### Author Declarations

In the present study, only anonymized datasets were accessed and used for the analysis after obtaining approval from the Qatar Biobank Institutional Review Board (E/2017/QGP-RES-PUB-008/0014).

